# Pharmacokinetics and Pharmacodynamics of (Val)Ganciclovir in Infants with Congenital Cytomegalovirus

**DOI:** 10.64898/2026.05.12.26353043

**Authors:** Brent Lindquist-Kleissler, Peter Kfoury, Jordan Stout, Ashlea Wilkes, Mark R. Schleiss, Albert H. Park, Joseph E. Rower

**Affiliations:** Center for Human Toxicology, Dept. of Pharmacology and Toxicology, University of Utah, Salt Lake City, UT; Divisions of Otolaryngology and Surgery, Dept. of Pediatrics, University of Utah, Salt Lake City, UT; Division of Pediatric Infectious Diseases, University of Minnesota Medical School, Minneapolis, MN; Department of Pediatrics, University of Utah, Salt Lake City, UT

**Keywords:** ganciclovir, valganciclovir, congenital cytomegalovirus, pharmacokinetics, pharmacodynamics

## Abstract

Ganciclovir (GCV), and its orally available pro-drug valganciclovir (VGCV), are preferred therapies for treating congenital cytomegalovirus (cCMV), however, their use carries a significant risk of neutropenia for the child. This risk limits dosing and effectiveness of VGCV, particularly in the treatment of infants with cCMV infection, who are at increased risk for sensorineural hearing loss (SNHL). We hypothesized that an improved understanding of the pharmacokinetics (PK) and pharmacodynamics (PD) of VGCV in cCMV-infected infants at risk for SNHL would inform strategies for optimizing safe and effective VGCV dosing. Participants were enrolled in one of two clinical studies interrogating the PK, safety, and efficacy of VGCV treatment in cCMV-infected infants at risk for SNHL. GCV exhibited a short median half-life of 2.02 h and the median (range) area under the 24 h concentration-time curve (AUC_24_) was 60.8 (26.8, 99.4) μg*h/mL. An AUC_24_ > 70 μg*h/mL was associated with an elevated risk of neutropenia (Fisher’s Exact *p* = 0.029). No associations between GCV PK and hearing outcomes were observed. Taken together, these results indicate vast inter-individual variability in GCV PK that is associated with dose-related toxicity, supporting the need for individualized dosing in the cCMV-infected population.

## Introduction

Congenital CMV (cCMV) occurs in 1 in 200 newborns,^1^ representing the most common congenital infection in humans and the leading non-genetic cause of sensorineural hearing loss (SNHL).^2–4^ Ganciclovir (GCV), and its orally bioavailable pro-drug valganciclovir (VGCV), are guanosine nucleoside analogs (NA), potent inhibitors of viral replication, and are used as first-line options for cCMV treatment. As with other NA, GCV competitively inhibits viral DNA polymerase chain elongation following phosphorylation to ganciclovir-triphosphate (GCV-TP). Early treatment of symptomatic cCMV-infected infants who have disease involving the central nervous system (CNS) with six months of GCV or VGCV (abbreviated collectively as (V)GCV) has demonstrated modest improvements in hearing and developmental outcomes,^5–7^ and more recent studies demonstrate the utility of six weeks of VGCV in cCMV infants with isolated SNHL.^8^ However, the effectiveness of antiviral therapy in ameliorating sequelae in asymptomatic cCMV-infected infants is not yet understood.

Existing data on (V)GCV pharmacokinetics (PK) and pharmacodynamics (PD) are limited, particularly for infants with cCMV infection. GCV exhibits a half-life of between 2 and 6 hrs, with exposures that vary significantly between individuals.^9–13^ A plasma GCV area under the 24 h concentration-time curve (AUC_24_) of between 40 and 60 μg*h/mL has been suggested for efficacy (i.e. reduced CMV viremia) in adult transplant recipients, but this threshold has not been validated in children.^14, 15^ Current guidance for infants with symptomatic cCMV infection suggests dosing VGCV twice-daily at 16 mg/kg,^16, 17^ however, clinical trial data suggests only modest hearing outcome benefits and that nearly 20% of infants develop neutropenia from this dosing strategy.^7^ Moreover, despite the potential benefit of (V)GCV for preventing SNHL in infants with asymptomatic cCMV infection, there is yet to be conclusive clinical trial evidence to support the use or appropriate dosing of (V)GCV in this population.

ValEAR was a randomized, placebo-controlled trial conducted to determine the safety and efficacy of twice-daily VGCV treatment in infants with cCMV infection and isolated SNHL.^18^ Dried blood spot (DBS) samples were longitudinally collected during the trial to describe VGCV PK. The current manuscript utilizes data from the ValEAR trial, as well as data from an open-label VGCV PK/PD study, to determine VGCV PK and associate GCV exposures with hearing outcomes (efficacy) and neutropenia (toxicity), with the hypothesis that a well-defined understanding of VGCV PK/PD will enable individualized dosing strategies and optimized clinical care of infants with cCMV infection.

## Methods

### Study Design

Study participants were enrolled in one of two studies, both conducted within outpatient settings. The first study, the ValEAR trial, was a multi-center, randomized, placebo-controlled clinical trial of VGCV treatment in infants with positive CMV titers and isolated SNHL disease.^18^ Study procedures were conducted under a central institutional review board (IRB) approval issued by the combined University of Utah/Primary Children’s Hospital board (cIRB_00090760, NCT03107871) and in accordance with the guidelines set forth by the Declaration of Helsinki. Infants meeting inclusion criteria (supplementary material) were enrolled with parent permission and treated with 16 mg/kg VGCV twice daily for 6 months. Venipuncture blood for safety laboratory values (including absolute neutrophil count, ANC) was collected at baseline, week 2, week 4, and monthly thereafter until month 7 after randomization. Additionally, these blood samples were used to prepare up to 5 x 30 µL DBS per time point for PK analyses. Auditory tests were conducted at baseline, 8, 14, and 20 months after randomization.

The second study was an open-label PK study of children receiving 16 mg/kg VGCV twice daily to treat moderate to severe CMV disease (either suspected or confirmed to be cCMV) at Primary Children’s Hospital in Salt Lake City, UT (IRB_00113183). Parental permission was obtained for participants who met the inclusion criteria (CMV infection and undergoing VGCV treatment, < 17 years of age). In addition to blood samples for PK, these participants had standard-of-care safety laboratory blood draws and auditory testing per clinical protocols. PK sample collection occurred at the participant’s clinical visits, at approximately 2-week intervals, and targeted samples collected prior to dose and at 30 min, 1 h, 2 h, 4 h, or 8 h post-dose. Blood for PK was collected via venipuncture and used to prepare up to 5 x 30 µL DBS per time point. DBS samples from both studies were allowed to dry for 2-4 h prior to storage at –80 °C. PD outcomes (ANC values and auditory test results) were obtained by chart review.

### GCV Bioanalysis

DBS GCV concentrations were quantified using a previously published, validated ultra-high-performance liquid chromatography-tandem mass spectrometry (LC-MS/MS) method.^19^ For each sample type (calibrators, quality controls, and unknowns), a 6 mm Harris Uni-Core^TM^ punch was taken from the center of the DBS and transferred to an appropriately labeled 13 x 100 mm borosilicate tube, followed by addition of an internal standard (GCV-d5). GCV was eluted from the DBS punch with methanol (MeOH) and sonication, followed by sample centrifugation. The supernatant was transferred to a fresh tube, then dried and reconstituted in ultrapure water (UPH_2_O) before injecting 30 μL for analysis. Analysis was conducted on a Thermo Scientific TSQ Vantage MS/MS coupled to a Thermo Scientific Accela UHPLC pump and autosampler. A Phenomenex® Synergi^TM^ 2.5 µm Polar-RP (100 x 2 mm) analytical and guard column, along with an isocratic mobile phase of 0.1% formic acid in water (99%) and MeOH (1%) delivered at a flow rate of 0.250 mL/min were used to achieve chromatographic separation. The MS/MS was operated in positive electrospray ionization (ESI) mode, monitoring mass transitions of 256.1->152.1 and 261.2->152.1 for GCV and GCV-d5, respectively. Samples were quantified using a calibration curve fit with a 1/x weighted linear regression between 10 and 10,000 ng/mL.

### Real-Time CMV PCR Assay

A sample consisting of 3 x 3 mm DBS punches for all collected samples was sent to the Department of Pediatrics at the University of Minnesota for quantitation of CMV viremia using a well-described polymerase chain reaction (PCR) assay.^20^ Multiplex qPCR was performed on sample eluate as previously described^20^ in 25 μL total volume with 10 μL of template using LightCycler 480 Probes Master Mix (Roche, Basel, Switzerland) containing FastStart Taq DNA Polymerase, reaction buffer, dNTPs mix (with dUTP instead of dTTP), and MgCl_2_; as well as 0.4 μM primers, 0.1 μM probes, and 0.4 U/μL of uracil-DNA glycosylase (UNG). PCR was performed using the Lightcycler 480 (Roche) under these conditions: 40 °C for 10 min, 95 °C for 10 min, followed by 45 cycles of 95 °C for 10 s and 60 °C for 45 s, then a final hold step at 40 °C. The primers used for UL83 were: forward primer, GGACACAACACCGTAAAGC; reverse primer, GTCAGCGTTCGTGTTTCCCA; and probe, CFR610-CCCGCAACCCGCAACCCTTCAT-BHQ2. The primers used for NRAS (housekeeping gene) were: forward primer, GCCAACAAGGACAGTTGATACAAA; reverse primer, GGCTGAGGTTTCAATGAATGGAA; and probe, FAM-ACAAGCCCACGAACTGGCCAAGA-BHQ1. A standard curve for the UL83 control was generated using 10-fold dilutions from 10^6^ to 10 copies/µL of a plasmid (UL83 fragment cloned in pCR2.1, using primers UL83_TM857F and UL83_TM1138R). A standard curve for NRAS was generated using five 10-fold dilutions starting with 200,000 to 20 pg/µL of human genomic DNA. (Roche). Final results were expressed as genome copies per microgram of cellular DNA.

### Absolute Neutrophil Counts

ANC values were determined by the clinical laboratory at the participant’s enrollment site. For this analysis, neutropenia was defined as moderate (grade 3) or severe (grade 4) disease, defined by an ANC < 1000 neutrophils/µL.

### Audiologic Assessments

Unaided audiologic assessments were conducted by experienced pediatric audiologists and included auditory brainstem response (ABR, typically for those younger than 6 months of age or when individual ear thresholds could not be obtained via behavioral testing), behavioral audiometry (0.25 to 8 kHz, typically for those older than 6 months of age), distortion product otoacoustic emissions, and tympanometry.^21, 22^ ABR testing included click and frequency-specific tone-burst stimuli with correction factors aligned with behavioral thresholds.^23^ Hearing threshold results obtained when participants had a conductive component to their hearing loss were excluded.

For ABR assessments, hearing loss was defined as threshold responses >25 dB eHL for clicks or any of the tone-burst frequencies at 1, 2, or 4 kHz.^24, 25^ For behavioral testing, hearing loss was defined as any threshold >20 dB HL at 1, 2, or 4 kHz. Hearing change and progression were analyzed from at least 2 separate hearing assessments. Any thresholds noted below 20 dB for behavioral testing and below 25 dB for ABR testing were corrected to 20 dB before analysis, so only changes beyond normal hearing would be clinically significant. Pure-tone averages (PTA) were calculated using air-conduction thresholds at 0.5, 1, 2, and 4 kHz. When thresholds at all four frequencies were available, the PTA was defined as the arithmetic mean of these four values. In cases where data for one frequency were missing, the PTA was calculated as the mean of the remaining three frequencies. Using PTA, severity was defined as mild for 30 to 45 dB (ABR or 21 to 45 dB (behavioral), then for all testing methods; moderate for 46 to 70 dB, severe for 71 to 90 dB, and profound for >90 dB. This approach was used to maximize data retention while maintaining a representative measure of speech-relevant hearing sensitivity. A progressive hearing loss was defined as a greater than 10 dB change in a click ABR, greater than 15 dB in one frequency, or greater than 10 dB change in more than one frequency.

### PK/PD and Statistical Analyses

PK parameters were calculated in Phoenix WinNonLin v8.6.1.6 using a non-compartmental analysis (NCA) with the linear trapezoidal/linear interpolation method. As blood volume limitations prevented the collection of serial samples within a single day, concentrations from distinct days were combined to make a concentration-time curve for a single dose. The appropriateness of this assumption is supported by the short half-life of GCV in plasma,^12, 26^ which is expected to result in minimal accumulation of GCV due to complete drug elimination within each dosing interval. Parameters of specific interest from the NCA included half-life (t_1/2_) and AUC. AUC was initially calculated as AUC_12_ (representing the AUC across the dosing interval, or AUC_τ_), then doubled to obtain an AUC_24_ for comparison to prior descriptions of therapeutic exposure targets.

GraphPad Prism v10.6.1 was utilized for data visualization and statistical analyses. Associations between continuous variables were explored using Pearson’s rho correlation. A receiver operating characteristic (ROC) curve analysis was conducted to determine a hypothesized threshold for neutropenia risk. The risk for developing neutropenia at a given GCV exposure (as determined from the ROC curve analysis, i.e., GCV AUC_24_ > 70 μg*h/mL) was tested using Fisher’s exact test.

## Results

### Study Population

A total of 26 participants were enrolled in these studies. Participant demographics are presented in Table 1. Of the 10 participants in the ValEAR study, five received a placebo and thus did not have measurable GCV concentrations. One participant in the treatment arm of the ValEAR study had measurable GCV in only 2 of 8 collected DBS samples (and only 3 of 8 had measurable GCV-TP)^27^ and was excluded from data analysis due to likely being nonadherent. In the open-label PK study, two participants only had one sample collected and were not used for the data analysis. A final open-label PK study participant had a concentration-time profile inconsistent with other participants (i.e., a pre-dose concentration greater than the concentration observed at 4 h), indicative of errant recordings of time post dose, and so was excluded. The total sample population, therefore, consisted of 17 participants for PK analysis. A total of 154 samples were collected from all participants, of which 66 were from participants used in the PK analyses (∼4 per participant). The resulting concentration-time profile is shown in Figure 1.

**Figure 1.**
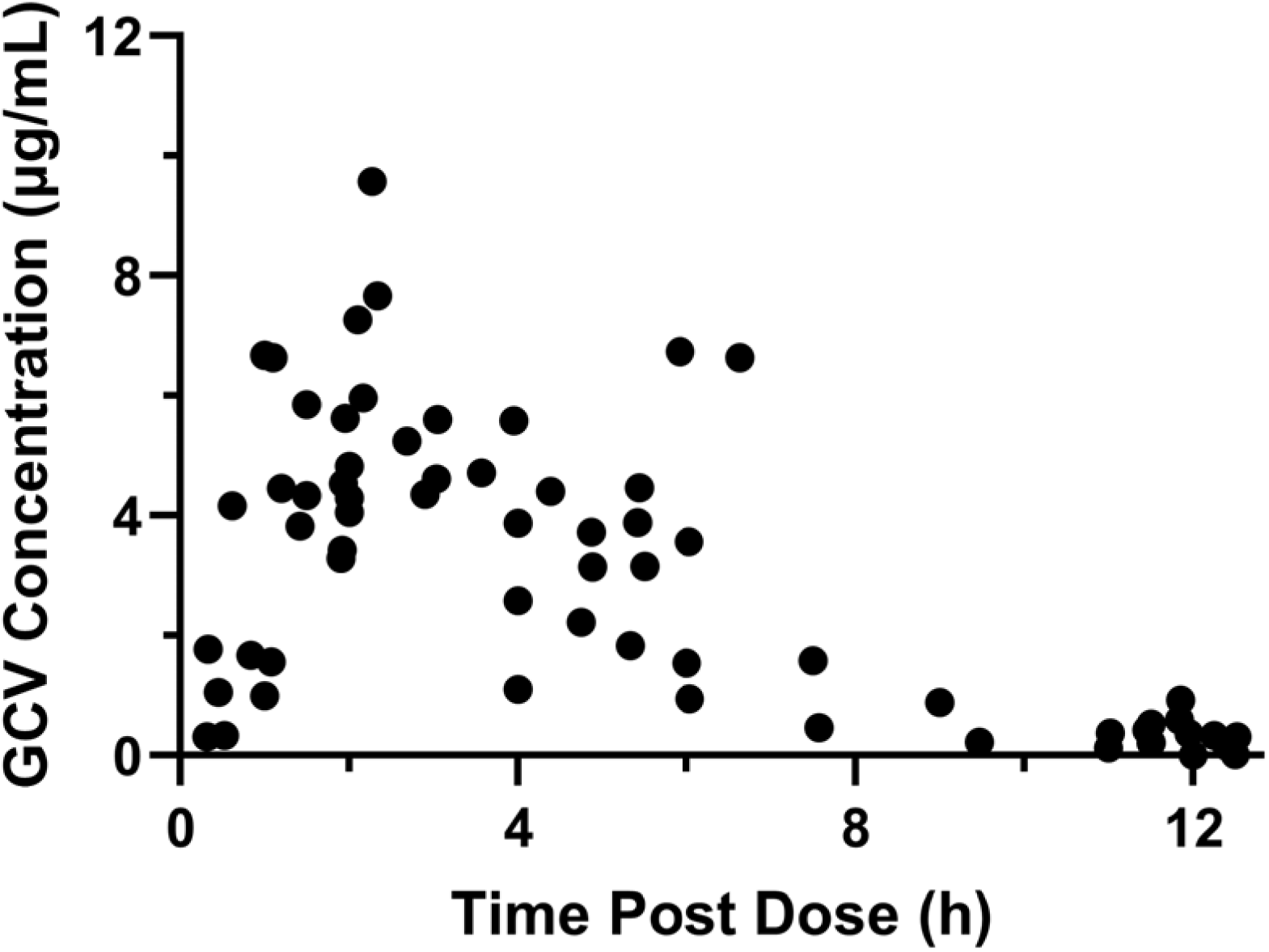
Concentration-time curve of GCV following VGCV administration. Each black dot is from a single DBS sample. The figure represents 66 samples from 17 participants (∼4 per participant).

**Table 1.** Participant demographics.

|  | ValEAR Study | Open-Label PK Study | Total |
| --- | --- | --- | --- |
| Participants (n) | 10<br>(5 treated, 5 placebo) | 16<br>(2 with $\leq 1$ DBS) | 26 |
| Sex | 6 Female, 4 Male | 9 Female, 7 Male | 15 Female, 11 Male |
| Race | 9 White, 1 Black | 13 White, 3<br>Unknown/Other | 22 White, 1 Black, 3<br>Unknown/Other |
| Weight (kg)<sup>1</sup> | 7.12 (6.04, 7.93) | 6.65 (4.95, 11.0) | 7.08 (5.10, 9.05) |
| Age (yr)<sup>1</sup> | 0.40 (0.30, 0.74) | 0.31 (0.18, 1.88) | 0.34 (0.20, 0.98) |
| Samples (n) | 63 | 57 | 120 |

<sup>1</sup>The continuous covariates weight and age are reported as median (interquartile range)

### PK Analysis

Summary results from the NCA are described in Table 2 for the 17 participants with sufficient data for PK analysis. Due to the expected short half-life of GCV,^12, 26^ we assumed complete elimination of GCV during the dosing interval. This assumption has several key benefits for our analysis, as it suggests that there is no accumulation of GCV across multiple doses. This allows the combination of data from across several doses as if they represented a single dose and allows the calculation of AUC_24_ as double the calculated AUC_12_ (which represents AUC_τ_). The median (interquartile range, IQR) GCV half-life determined in the current study was 2.02 (1.55, 2.77) h, supporting this assumption. The median (IQR) AUC_12_ was 30.4 (20.5, 34.0) µg*h/mL, yielding a median (IQR) AUC_24_ of 60.8 (IQR 40.9, 68.0) µg*h/mL. AUC_24_ values > 60 μg*h/mL occurred in ∼50% of participants (n=9, Figure 2), with two of these participants having an AUC_24_ > 90 μg*h/mL. The remaining participants were evenly split between AUC_24_ values within and below the hypothesized 40-60 μg*h/mL therapeutic target. The median T_max_ was 2.10 (IQR 1.50, 3.98) h and the C_max_ was 5.61 (IQR 4.31, 6.70) µg/mL. The median C_last_ was 0.333 (IQR 0.184, 0.519) µg/mL.

**Figure 2.**
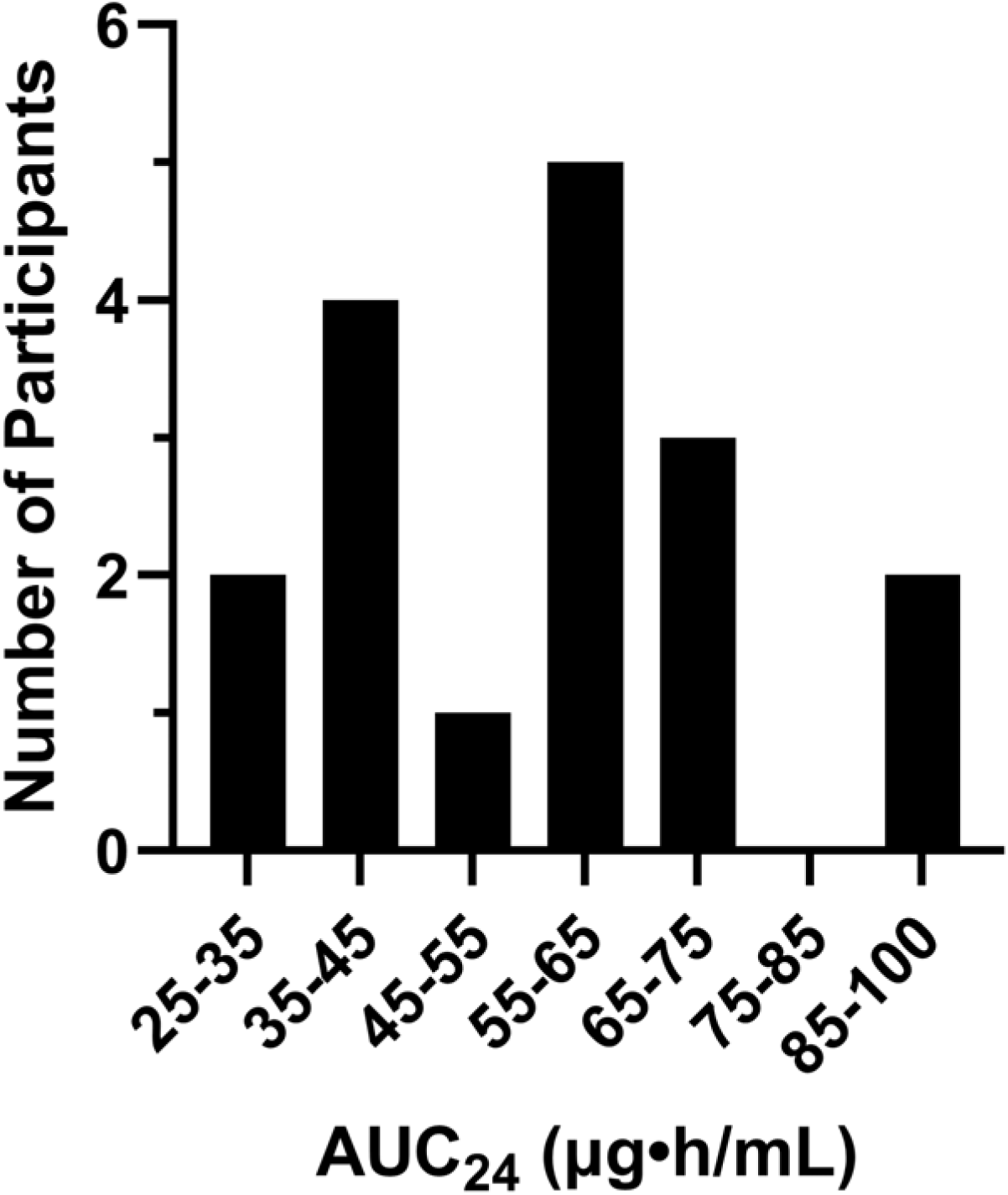
Distribution of AUC_24_ and number of participants in that range.

**Table 2.**
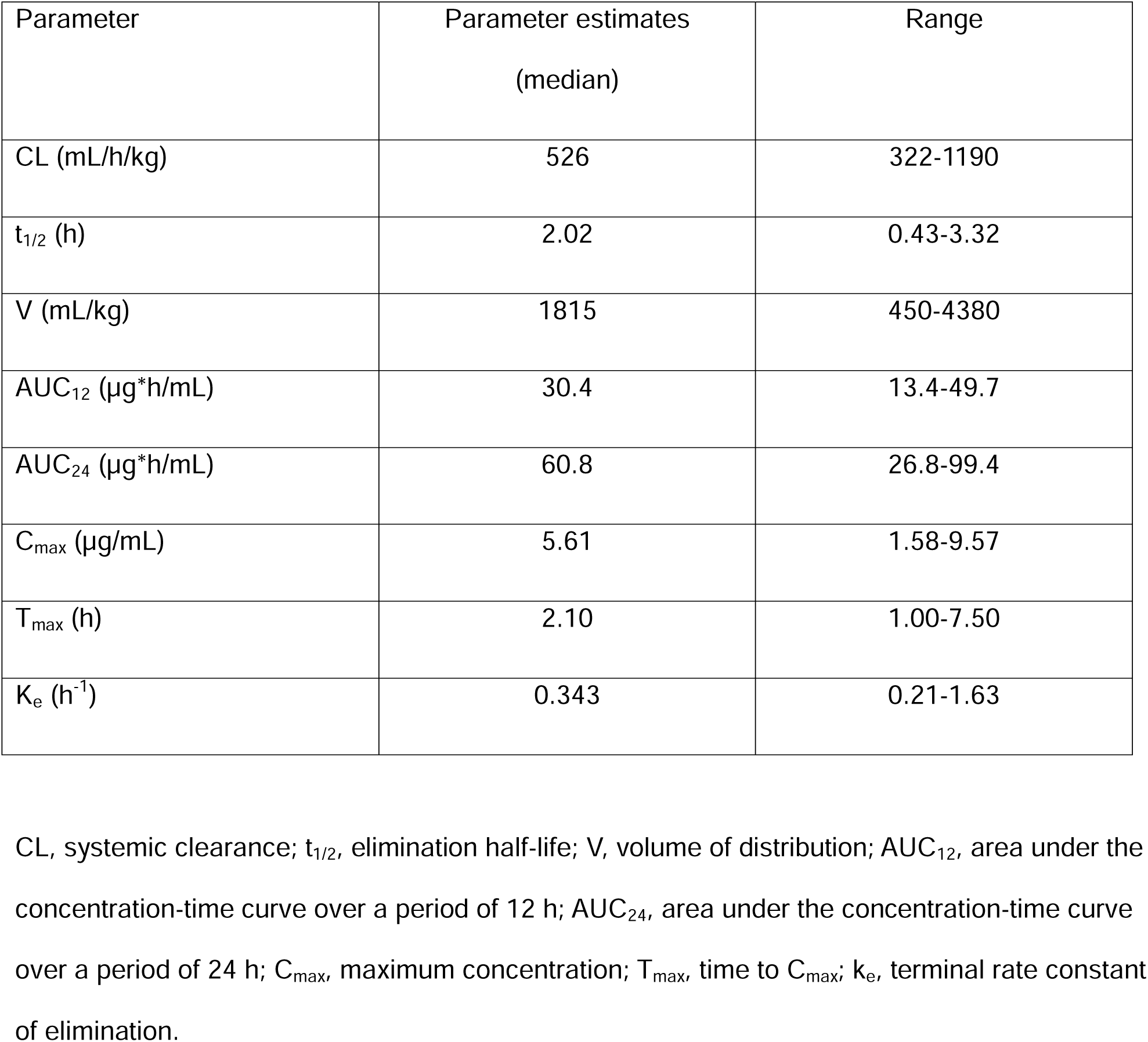
NCA pharmacokinetics parameters following VGCV administration.

| Parameter | Parameter estimates<br>(median) | Range |
| --- | --- | --- |
| CL (mL/h/kg) | 526 | 322-1190 |
| $t_{1/2}$ (h) | 2.02 | 0.43-3.32 |
| V (mL/kg) | 1815 | 450-4380 |
| AUC<sub>12</sub> (µg*h/mL) | 30.4 | 13.4-49.7 |
| AUC<sub>24</sub> (µg*h/mL) | 60.8 | 26.8-99.4 |
| C<sub>max</sub> (µg/mL) | 5.61 | 1.58-9.57 |
| T<sub>max</sub> (h) | 2.10 | 1.00-7.50 |
| K<sub>e</sub> (h<sup>-1</sup>) | 0.343 | 0.21-1.63 |

CL, systemic clearance; $t_{1/2}$ , elimination half-life; V, volume of distribution; AUC<sub>12</sub>, area under the concentration-time curve over a period of 12 h; AUC<sub>24</sub>, area under the concentration-time curve over a period of 24 h; C<sub>max</sub>, maximum concentration; T<sub>max</sub>, time to C<sub>max</sub>; k<sub>e</sub>, terminal rate constant of elimination.

### Neutropenia and AUC

An ROC curve analysis indicated a plasma GCV AUC_24_ > 70 µg*h/mL provided >80% sensitivity and specificity for predicting neutropenia risk. All 17 participants within the PK analysis had longitudinal ANC data for evaluation. Of these participants, 14 had AUC_24_ < 70 µg*h/mL, of whom 3 developed neutropenia. Three participants with AUC_24_ > 70 µg*h/mL were all neutropenic (Table 3). A Fisher’s exact test indicated an increased risk for neutropenia in those individuals with AUC_24_ > 70 μg*h/mL (*p* = 0.029). One of the 5 participants receiving placebo had consistent ANC values indicative of neutropenia throughout the study period, while the other four in the placebo arm remained at steady ANC values above the 1000 neutrophil/μL cutoff.

**Table 3.** Contingency table for Fisher’s exact test of AUC24 versus ANC levels.

|  | AUC<sub>24</sub> < 70 µg*h/mL | AUC<sub>24</sub> > 70 µg*h/mL |
| --- | --- | --- |
| Neutropenic | 3 | 3 |
| Non-neutropenic | 11 | 0 |

### CMV Viremia

A total of 100 DBS samples were available to measure CMV viremia across all but one participant (n=25 participants in total). Only 4 samples had sufficient viremia for amplification. Of these 4 samples, 2 were from a single ValEAR participant in the treatment arm of the trial, representing samples from baseline and 1 month after treatment cessation, suggesting an immediate rebound post-cessation of therapy. The third sample was from a participant in the treatment arm of the ValEAR trial at month 2 of treatment, however, this participant had only 2 of 8 collected samples with quantifiable GCV concentrations, indicative of likely treatment non-adherence.^27^ The final sample was collected approximately 2 weeks after treatment initiation and was the only sample collected from a child in the open-label PK study.

### Hearing Outcomes

Of the 22 participants either placebo-treated (n=5) or included in the PK analysis (n=17), three were excluded for the absence of a definitive cCMV diagnosis in their chart, all in the open-label PK study and all age >18 months. An additional 3 participants (2 placebo treated ValEAR participants, and one open label PK study participant) had only one auditory testing result during the study period for the best ear and were removed from the analysis, yielding a total of 16 participants for the per protocol best ear analysis (best ear was determined as the ear with the least SNHL). Prior to treatment initiation, 10 participants had normal hearing, 1 had mild SNHL, 2 had moderate SNHL, 2 had severe SNHL, and 1 had profound SNHL in the best ear. After treatment initiation, with an average follow-up of 13.2 months (SD= 7.2 months), 13 participants demonstrated normal hearing while 3 had severe SNHL (Figure 3a). Of the 4 participants with improved hearing, all participated in the ValEAR study: two had baseline moderate hearing that was measured as normal at the end of the study (one with AUC_24_=38.5 μg*h/mL and the other placebo-treated), one had profound hearing loss at baseline that measured as severe (AUC_24_=62.3 μg*h/mL), and the final participant had mild baseline hearing that measured normal at the study’s conclusion (AUC_24_=62.7 μg*h/mL). No clear association between GCV exposure and hearing outcome was identified within the best ear.

**Figure 3.**
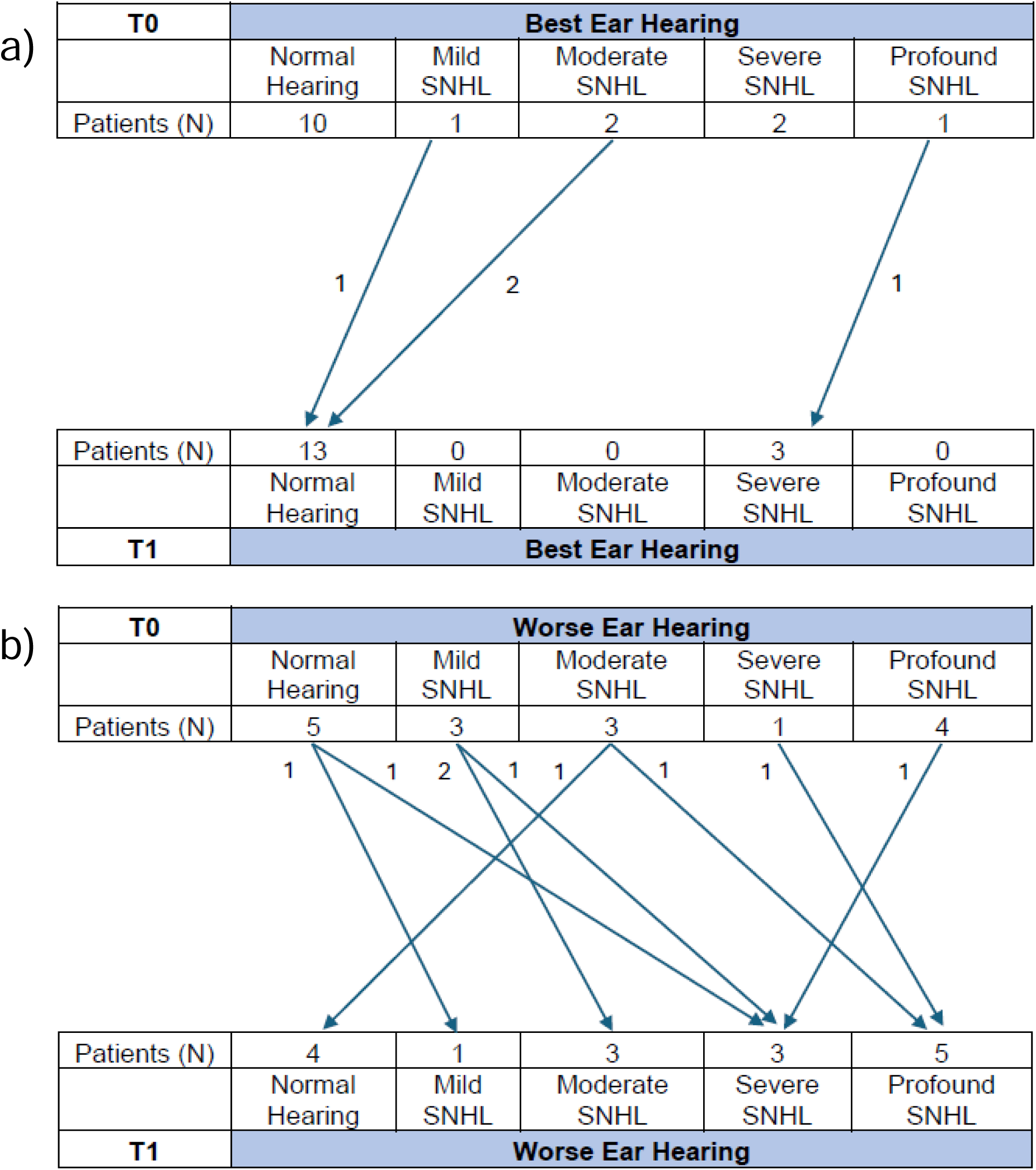
Hearing outcomes for N=16 participants at treatment initiation (T0) and at the last follow-up (T1). a) best ear hearing outcomes. b) worse ear hearing outcomes.

A secondary endpoint was to analyze worse ear hearing loss and utilized the same 16 participants described in the best ear analysis. Prior to treatment initiation, 5 participants had normal hearing, 3 had mild SNHL, 3 had moderate SNHL, 1 had severe SNHL, and 4 had profound SNHL in their worse ear. After treatment initiation, with an average follow-up of 13.2 months (SD= 7.2 months), 4 participants demonstrated normal hearing, 1 had mild SNHL, 3 had moderate SNHL, 3 had severe SNHL, 5 had profound SNHL (Figure 3b). Three individuals with the same or better hearing in their worse ear (n=9) received placebo; no individuals with worsening hearing (n=7) in their worse ear received placebo. Among individuals with measurable AUC_24_ values (i.e. excluding three participants administered placebo), the median AUC_24_ (IQR) was 63.3 (47.4, 71.8) µg*h/mL in participants with worsening hearing (n=7), compared to a median AUC_24_ (IQR) of 52.3 (42.2, 71.4) µg*h/mL in individuals (n=6) with improved or similar hearing in their worse ear (Mann Whitney *p* = 0.295).

## Discussion

Congenital CMV is the leading, nonhereditary cause of SNHL; however, due to the potential side effects (e.g., neutropenia), the utility of (V)GCV treatment remains unknown for cCMV-infected infants with isolated SNHL. With better understanding of (V)GCV pharmacology, the effectiveness of (V)GCV treatment for cCMV infection with isolated SNHL could be improved while limiting the risks of side effects like neutropenia. This has the vast potential to increase therapeutic success and potentially decrease the rates of SNHL within this population.

Overall, our NCA analysis yielded PK results in good alignment with previously published data. Based on our NCA calculations, the median (range) AUC_24_ for our two combined studies was 60.8 (26.8, 99.4) µg*h/mL. One prior analysis in a similar cohort receiving 16 mg/kg oral VGCV observed a mean AUC_24_ of 54.0 µg*h/mL (AUC_12_ = 27.0 µg*h/mL).^10^ Other work administering either 4 or 6 mg/kg GCV (expected to yield similar exposures to a 16 mg/kg oral VGCV dose) found mean AUC_24_ values to be 53.4 µg*h/mL (AUC_12_ = 26.7 ± 3.6 µg*h/mL) and 64.6 µg*h/mL (AUC_12_ = 32.3 ± 3.8 µg*h/mL), respectively.^10, 26^ While the median AUC_24_ in the current study is similar to previous studies, our data indicate a high level of inter-individual variability of GCV exposure. Indeed, one individual in our study had an AUC_24_ of 26.8 μg*h/mL, approximately two-fold lower than the target therapeutic exposure, while two others had AUC_24_ near 100 μg*h/mL, approximately two-fold higher. The extensive inter-individual variability supports the need for individually optimized VGCV dosing, as the current one-size-fits-all approach does not appear sufficient to ensure safe and efficacious therapy. One covariate to consider using in individualized dosing is renal function, which, unfortunately, was not available in our dataset. Ganciclovir is primarily excreted from the kidney; thus, impaired renal function is expected to impact GCV PK. Indeed, literature suggests lowering GCV dosing for participants with mild renal impairment, though it is unclear how widely applied this is within clinical practice.^28^

Of the 17 participants included in the PK analyses, 9 (52.9%) had AUC_24_ values above the recommended range of 40-60 µg*h/mL, 4 participants (23.5%) were below the recommended range, and 4 participants (23.5%) were within the recommended range. Prior reports of GCV PK have predicted 21% of children attain GCV exposures within the hypothesized exposure target of 40 to 60 μg*h/mL, in line with the findings of the current study.^29^ Seven of 22 participants, 32% of the study population, developed neutropenia, including all participants with a GCV AUC_24_ > 70 μg*h/mL (n=3). Combined, these data indicate that the standard VGCV dosing regimen of 16 mg/kg twice daily frequently results in GCV exposures exceeding the hypothesized target efficacy threshold, placing the child at an elevated risk for neutropenia. As a result, VGCV effectiveness, particularly in the setting of cCMV disease, may benefit from reduced dose and/or frequency of VGCV administration.

In one previous study, symptomatic infants were treated with placebo or 6 mg/kg of GCV twice daily for 6 weeks. Of the treated participants, 84% had improved hearing or experienced no change in best ear hearing, while none had worsening hearing from baseline.^5^ Another study compared hearing outcomes between symptomatic infants treated for 6 weeks or 6 months with 16 mg/kg of VGCV. Of the participants in the 6-month group, 73% had best ear hearing that improved or remained normal after 12 months, compared to 57% in the 6-week group.^7^ A retrospective study on children with cCMV infection who were referred to an audiologic center for a 6-year follow-up, determined that only one child treated with VGCV experienced hearing deterioration.^30^ In our study, a total of 84 hearing assessments on 22 infants were conducted; however, we were unable to determine any PK/PD trends, likely due to the lack of variability within individuals’ hearing outcomes. One potential factor is that adverse events associated with high GCV AUC_24_ values may lead participants to become non-adherent or even stop using (V)GCV.^17, 27^ The population from the other studies were younger than ours, which could be one of the factors leading to overall better hearing outcomes, as GCV has been described to better penetrate the inner ear in younger mice.^31^ Another difference is that our study focused on treating cCMV-infected infants with isolated SNHL, whereas the other study focused on treating symptomatic infants; it is possible that disease severity alters treatment outcomes. Of note, no participant had worsened hearing in their best ear, while worsening hearing occurred in 47% (n=8 out of 17) of participants’ worse ears. This supports the importance of testing hearing thresholds for both ears, as noted by a previous study.^32^ Further work evaluating hearing outcomes in a larger cohort of cCMV-infected infants with or without isolated SNHL is needed to determine the benefit of prophylactic therapy in this population.

Our NCA calculations yielded a GCV half-life of ∼2 h in plasma, in agreement with previously published data.^10, 12, 26^ However, as with other NA, GCV requires sequential phosphorylation to the GCV-TP moiety. Substantial data exists for other NA that plasma NA half-lives are shorter than their intracellular form, due to ion-trapping of the TP form within the cell.^33^ Thus, it is reasonable to assume that GCV-TP may accumulate within cells to an extent greater than would be predicted from its plasma half-life, and that this accumulation will be cell-specific. Indeed, recent work using the samples described in the current study found that GCV-TP has a half-life of ∼21 days in DBS, which represents red blood cell concentrations.^27^ It is well documented that ganciclovir accumulation leads to bone marrow toxicity, where neutrophils originate, supporting a critical need to better understand the cellular kinetics of GCV-TP to refine dosing strategies.^34, 35^ Indeed, monitoring GCV-TP, rather than the parent GCV, may provide more accurate insight into an infant’s risk of neutropenia and allow more personalized dosing to target the therapeutic window.

Although the current study generated interesting findings, it also has limitations that warrant acknowledgment. Though the current study population is small, it is among the largest focusing on VGCV treatment in a pediatric population with cCMV. The largest study enrolled 96 participants,^7^ followed by our study enrolling 26 participants. There are two slightly larger studies that each enrolled 27 participants with cCMV, however they were treated with GCV and not VGCV.^26, 36^ Notably, this current study focused on directly relating PK to viremia, neutropenia, and hearing outcomes within the same population, while the previously mentioned three largest studies only looked at PK or PD. Additionally, previous studies focused on symptomatic cohorts, while the ValEAR trial focused on asymptomatic infants and infants with isolated SNHL. These groups are typically not treated due to potential side effects (e.g. neutropenia) which are especially dangerous to these non-immunocompetent populations, that nevertheless can experience life-long hearing loss. Better understanding of the PK/PD of (V)GCV treatment in these populations is vital for improving long term outcomes. The absence of robust serum creatinine sampling within our study population prevented the evaluation of creatinine clearance as a potential covariate informing variability in plasma GCV PK. The study also had sparse sampling and missing data, which may have limited the identification of PK/PD relationships, particularly with respect to hearing outcomes. Future work can address these concerns by conducting a larger study that includes more frequent sampling, more robust and directed data collection, and the enrollment of a more diverse study population.

## Conclusions

The GCV exposures determined in the current study exhibit high inter-individual variability, with more participants falling outside the hypothesized target exposure than within. A GCV AUC_24_ of 70 µg*h/mL was identified as a threshold for increased risk of neutropenia. These findings demonstrate the need for a better understanding of GCV pharmacology in infants, including analysis of the active GCV-TP moiety, to enable refined and personalized dosing strategies in children with cCMV infection.

## Data Availability

Data that can be reasonably shared without compromising a participant's identity can be made available upon request to the corresponding author.

## Acknowledgments

The authors are grateful to the study participants and their parents who volunteered to participate in the study. Technical support for PCR assay development from Mark Blackstad (University of Minnesota Department of Pediatrics) is acknowledged.

## Author Contributions

BLK contributed to data analysis and manuscript writing/editing. PK, JS, and AW facilitated sample collection, assisted with data generation, and reviewed the manuscript. AHP received funding for the study and reviewed the manuscript. JER contributed to data generation/analysis, manuscript writing/editing, and received funding for the project. MRS contributed to data generation/analysis and manuscript writing/editing.

## Ethics Approval

All study procedures were approved by the University of Utah and Primary Children’s Hospital IRB (IRB_00113183 and IRB_00090760) and conducted in accordance with the ethical principles in the Declaration of Helsinki. Parental permission was obtained for all participants in the study.

